# Additive pre-diagnostic and diagnostic value of routine blood-based biomarkers in the detection of colorectal cancer in the UK Biobank cohort

**DOI:** 10.1101/2022.11.10.22282166

**Authors:** Gizem Tanriver, Ece Kocagoncu

## Abstract

**Background:** Survival rates from colorectal cancer (CRC) are drastically higher if the disease is detected and treated earlier. Current screening guidelines involve stool-based tests and colonoscopies, whose acceptability and uptake remains low. Routinely collected blood-based biomarkers may offer a low-cost alternative or aid for detecting CRC.

**Methods:** Here we aimed to evaluate the pre-diagnostic and diagnostic value of a wide-range of multimodal biomarkers in the UK Biobank dataset, including sociodemographic, lifestyle, medical, physical, and blood and urine-based measures in detecting CRC. We performed a Cox proportional hazard and a tree-boosting model alongside feature selection methods to determine optimal combination of biomarkers.

**Results:** In addition to the modifiable lifestyle factors of obesity, alcohol consumption and cardiovascular health, we showed that blood-based biomarkers that capture the immune response, lipid profile, liver and kidney function are associated with CRC risk. Following feature selection, the final Cox and tree-boosting models achieved a C-index of 0.67 and an AUC of 0.76 respectively.

**Conclusions:** We show that blood-based biomarkers collected in routine examinations are sensitive to preclinical and clinical CRC. They may provide an additive value and improve diagnostic accuracy of current screening tools at no additional cost and help reduce burden on the healthcare system.

## Background

Colorectal cancer (CRC) is the fourth most common type of cancer after breast, prostate, and lung cancer in England, constituting 11.4% of all cancer cases [1]. Despite falling mortality rates over the last decades, CRC remains the second most common cause of cancer-related deaths with an age-standardised overall survival rate of 78% at year 1, subsequently falling to 59% after 5-years [2,3]. Metastasis is the main cause of mortality, observed in half of the CRC patients, with liver as the most common distant site [4]. Early detection drastically improves surgery and treatment outcome. Nearly 90% of patients survive at 5-years post-diagnosis, compared to only 10% in advanced-stages [5]. Further, early detection can reduce treatment costs and save healthcare professionals’ valuable time [6]. Therefore, there is an urgent need for biomarkers that are sensitive and specific to early stages of CRC, well-accepted by patients, and scalable to the national level at lower costs.

Current gold standards of early diagnostic tools are faecal immunochemical test (FIT; 79% sensitivity, 94% specificity) in the UK and multitarget stool DNA test in the US (MT-sDNA, Cologuard^®^; 92% sensitivity, 86.6% specificity) [7,8], which are followed up by colonoscopy and polypectomy, if positive. While stool-based tests show good diagnostic performance, are non-invasive, safe and simple to perform, patient acceptability and uptake remains low [9]. Instead, blood-based biomarkers offer a more acceptable, low-cost alternative to stool-based tests [10], are routinely used across hospitals in the UK, and have the potential to be adopted for home-testing. Cancer-specific blood-based markers detect presence of genomic and epigenomic markers, circulating tumour cells, or specific protein markers such as carcinoembryonic antigen (CEA) [11]. Although these markers show diagnostic potential for CRC, they are not part of routine blood panels.

Numerous biochemical and hematological measures, tested in routine blood panels, show associations with the risk for developing CRC. Some of these biomarkers measure systemic inflammation, which include acute-phase proteins such as C-reactive protein and albumin [12] and white blood cells such as lymphocytes and neutrophils [13,14]. Others include liver enzymes [15,16] and lipids [17] as well as hematological measures such as hemoglobin and platelets which might indicate rectal bleeding and anemia as a result of CRC [13,18]. Individually these biomarkers are not highly sensitive or specific to CRC. When combined, however, they might have the potential to detect wider, multifaceted aspects of the molecular changes that occur in CRC. Identifying optimal combinations can be achieved by using statistical or machine learning (ML) models, alongside data-driven feature selection methods.

In this study, using the longitudinal UK Biobank dataset, we investigate the diagnostic potential of a wide range of biomarkers including medical, socioeconomic, and routinely collected blood and urine-based laboratory measures, in detecting preclinical and clinical CRC. Here we adopt a data-driven approach and perform (i) feature selection to identify biomarkers sensitive to CRC; (ii) a survival analysis to determine the optimal combination of pre-diagnostic biomarkers and quantify their contribution to the risk of developing CRC; and lastly (iii) a classification model to determine the combination of diagnostic features that can classify CRC cases from healthy participants with high accuracy. Survival analysis focused on incident cases of CRC and used Cox proportional hazards regression to model multivariate associations. The classification model used the GPBoost algorithm, which combines mixed effects models with tree-boosting, and focused on prevalent cases. With these two methods, we aimed to identify biomarker combinations sensitive to diagnostic and clinical CRC.

## Methods

### UK Biobank study

UK Biobank is a large scale, population-based cohort aged 40-69 years, recruited between 2006 and 2010 across 22 UK-based assessment centres, and aiming to follow up for 20 years [19]. The dataset comprises sociodemographic, psychosocial, lifestyle, family history, clinical, physical, cognitive, activity monitoring, biochemical, imaging, health linkage to a wide range of electronic healthcare records, and genomics data from over 500,000 participants.

At the baseline visit, a touchscreen questionnaire and a computer assisted interview were completed. Physical and functional measures, and blood, urine and saliva samples were collected. The baseline assessments were repeated on a smaller subset of the cohort (20,000-25,000 participants). The time lag between visits was approximately 4 years on average (range: 1-10 years).

All participants were registered in the UK National Health Service. Cancer outcomes were taken from electronic healthcare records, hospital episodes statistics, the National Cancer Registry, self-reports validated by the study nurse and death certification data. Outcomes were coded using the International Classification of Diseases 10^th^ revision (ICD-10) system.

### Participants

Initial sample size was 502,411 participants. 387,773 participants (77.2%) were healthy controls (HC), defined as those who have not received a cancer diagnosis before or during the study. To define the CRC group, we used ICD-10 codes for malignant neoplasm of colon including caecum, appendix, ascending colon, hepatic flexure, transverse colon, splenic flexure, descending colon, sigmoid colon, overlapping lesion of colon, and colon unspecified (C18.0-9), rectosigmoid junction (C19) and rectum (C20). There were 2,317 participants with CRC at baseline (0.46%), and 6,237 incident cases (1.24%) on follow-up visits. Among incident cases, 6,116 (98%) were diagnosed after their last study visit.

### Measures

We included 72 sociodemographic, physical, medical, lifestyle and biochemical measurements in our analysis. Cancer-related variables were only used to describe the patient population and were not used as predictors in the analyses.

#### Sociodemographic measures

age at screening visit, sex, ethnicity, Townsend deprivation index as an index of socioeconomic status, and highest education qualification.

#### Physical measures

height, body mass index (BMI), pulse, diastolic and systolic blood pressure (BP), waist-to-hip ratio, trunk-to-leg fat ratio, metabolic rate, impedance, grip strength of the dominant hand, and self-reported sleep duration in hours.

#### Medical history

Family history of cancer and CRC, disease history for inflammatory bowel disease (IBD), cardiovascular disease (CVD), liver and biliary disease, diabetes, self-reported overall health rating (on a scale of 1 to 4 from Excellent to Poor), and regular aspirin and statin use.

#### Lifestyle

Smoking and alcohol consumption frequency, intake frequency of oily fish, processed and red meat, summed metabolic equivalent task (MET) minutes per week for all activity based on International Physical Activity Questionnaire.

#### Full blood count (FBC) & Biochemistry

Biochemical measures were assessed in serum and urine. Serum markers consisted of white blood cell (WBC) count, red blood cell (RBC) count, hemoglobin concentration, hematocrit percentage, platelet count, lymphocyte percentage, apolipoprotein A and B, urea, cholesterol, C-reactive protein (CRP), cystatin C, high density lipoprotein, insulin-like growth factor 1 (IGF-1), low density lipoprotein, sex hormone binding globulin (SHBG), testosterone, total protein, triglycerides, vitamin D, mean corpuscular volume, percentages of monocytes, neutrophils, eosinophils, basophil, nucleated RBCs, and reticulocytes, albumin, alkaline phosphatase (ALP), alanine aminotransferase (ALT), aspartate aminotransferase (AST), direct bilirubin, calcium, gamma glutamyltransferase (GGT), glucose, glycated hemoglobin A1C (HbA1C), phosphate, total bilirubin and urate. An additional measure was included in the models as a covariate to indicate whether the participant fasted before the blood draw. Markers assessed in urine were creatinine, potassium, and sodium.

#### Cancer related variables

Age at cancer diagnosis, cancer site, behaviour of the cancer tumour, distinct diagnoses of cancer, and whether the participant had been previously screened for CRC.

### Pre-processing

All preprocessing and data analysis steps (**Fig 1**) were carried out with Python 3. Participants whose consent date was not available (*N* = 2), who withdrew consent (*N* = 158), who had other primary cancers (*N* = 105,924) or concurrent cancers (*N =* 1,458) were excluded. We removed outliers outside the 0.1^th^ and 0.99^th^ percentiles. To simplify interpretation of results, ethnicity was re-coded into white and non-white, and education level as university and non-university. Summed MET minutes was binned into 5 quintiles. To maximise available data in the following analyses, we opted for methods that can handle sparse data when possible, and coded missing data in categorical measures as an ‘unknown’ category. We performed group comparisons on baseline data using two-tailed chi-square tests and between samples t-tests and corrected for multiple comparisons using the false discovery rate. Analysis-specific pre- processing steps are given in respective sections.

**Fig 1.**
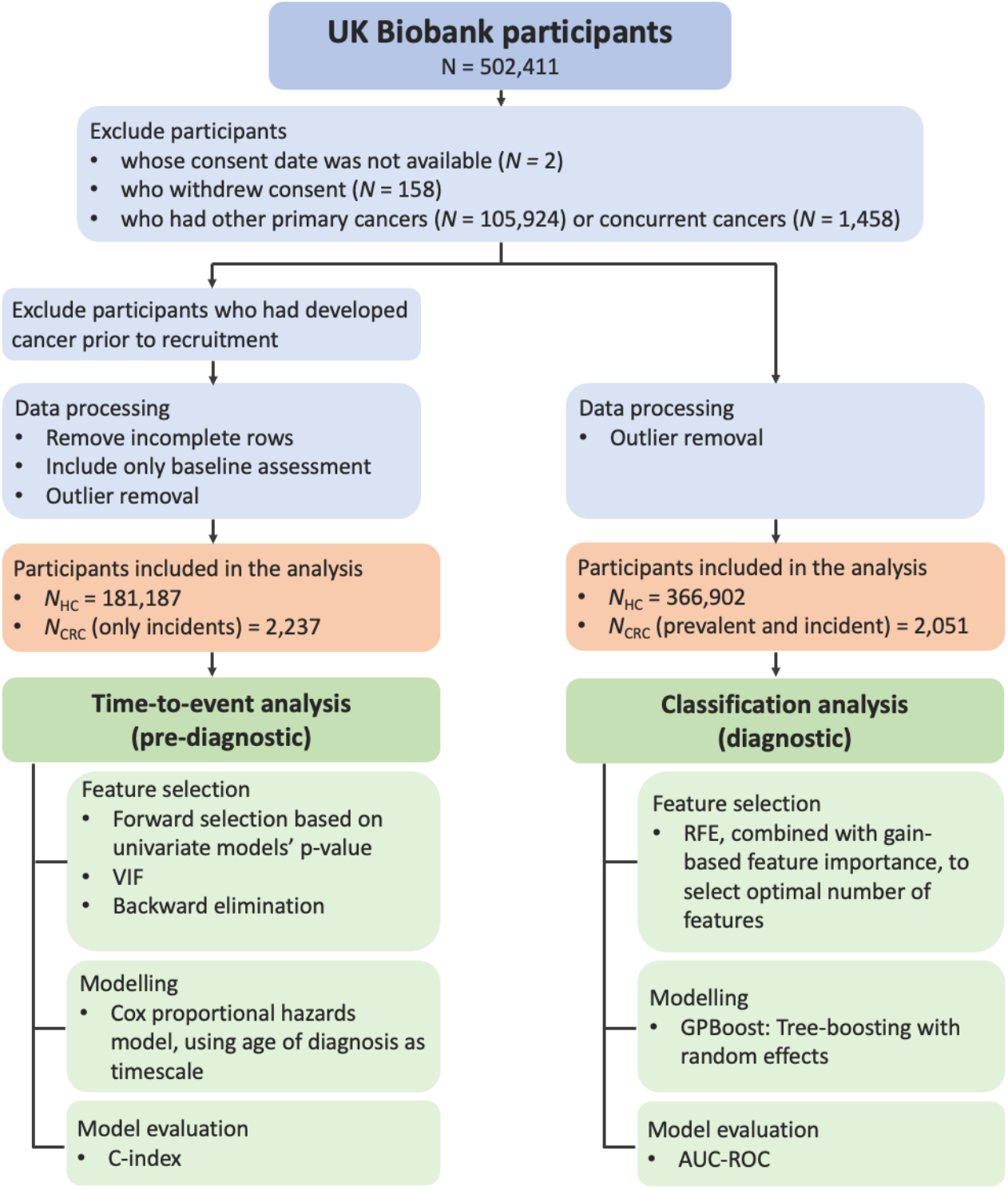
Analysis pipeline showing the distinct feature selection, modelling, and model evaluation steps of the time-to- event and classification analyses that aimed to identify pre-diagnostic and diagnostic biomarkers sensitive to CRC.

### Time-to-event analysis

We used Cox regression model to assess the effect of the above biomarkers on the age of diagnosis and associated risk for CRC using the Lifelines package [20]. In addition to the filtering steps explained in pre- processing, we removed participants who developed any cancer prior to recruitment (*N =* 6,740). All participants were followed until the censoring date, 29th February 2020. We calculated survival based on participant’s age. Covariates used in the model were measures from the initial screening visit. Since Cox regression does not handle missing data, rows with missing values were removed.

A data-driven approach was adopted to find the optimal set of covariates to model in Cox regression. We first split the dataset into 80% training and 20% test sets, stratified by label. We ran a forward feature selection, where each covariate was univariately fitted to the dataset, and any covariates that exceeded *p >* 0.10 were removed from the list of features. We then tested for multicollinearity in the dataset by calculating variance inflation factors (VIF) and removed any covariate exceeding *VIF* > 10. Finally, we used backward feature elimination where we initially modelled all the remaining variables, then iteratively removed the variable with highest non-significance at *P >* 0.05, until all covariates in the model were significant. The final model was then evaluated with the test set. We report the concordance index (C-index) for both training and test sets, as an index of model’s performance.

### Classification analysis

We used tree-boosting with grouped random effects model to classify CRC vs HC using both baseline and repeat visits (where available). We used Python implementation of *GPBoost* which combines powerful tree-boosting algorithms with mixed effects models – which are commonly used when working with grouped data [21,22]. The tree-boosting part utilises *LightGBM*, a highly efficient type of gradient boosting decision tree, which handles missing values and works with categorical variables [23].

Here we used the same feature processing and outlier removal procedures as before. Unlike the survival analysis, we included both prevalent and incident cases, and retained incomplete rows. We assigned a binary label for CRC to each study visit of a participant based on whether they had developed CRC at the time of the visit. We first split the dataset into 80% training and 20% test sets, and then split the training set into 5 cross-validation (CV) folds for feature selection experiments. We evaluated the model performance using Area Under the Receiver Operating Characteristic Curve (AUC-ROC).

To determine the number of features, we implemented Recursive Feature Elimination (RFE) [24]. RFE is a wrapper type of feature selection algorithm, which recursively prunes the least ‘important’ features until the desired number of features is reached. We performed a grid search with 1-5 features, using ‘gain’- based feature importance provided by our model. The optimal number of features (N_features_) was then selected as the number of features which achieved the highest score in 5-fold CV. We also calculated feature importance values for each split, normalised by the total importance, and aggregated by mean across splits. We generated partial dependence plots (PDP) to assess the dependency of predicted CRC probability on each feature.

## Results

### Sample characteristics at baseline

Group comparison results of sociodemographic, medical history and key lifestyle measures are given in **Table 1**. Across sociodemographic measures, two groups had significantly different ages and sex. CRC group was significantly older and had a higher percentage of males than the controls. There were no significant differences on ethnicity, education level and socioeconomic status measured by the Townsend Deprivation Index. The majority of the participants on both groups were white and had tertiary education as their highest qualification. Both groups had a negative mean score of Townsend Deprivation Index, indicating relative affluence in each group relative to the general population.

**Table 1.**
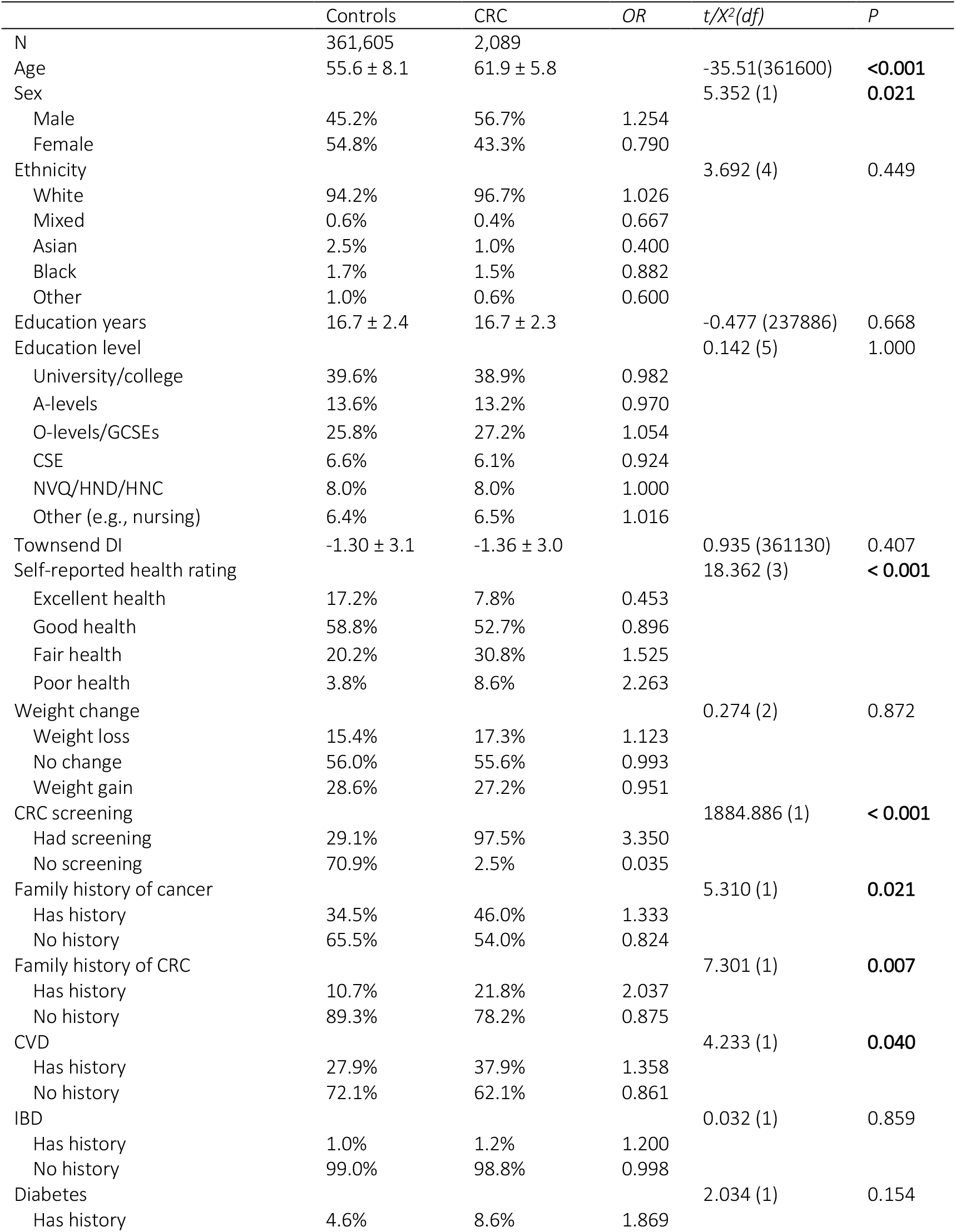
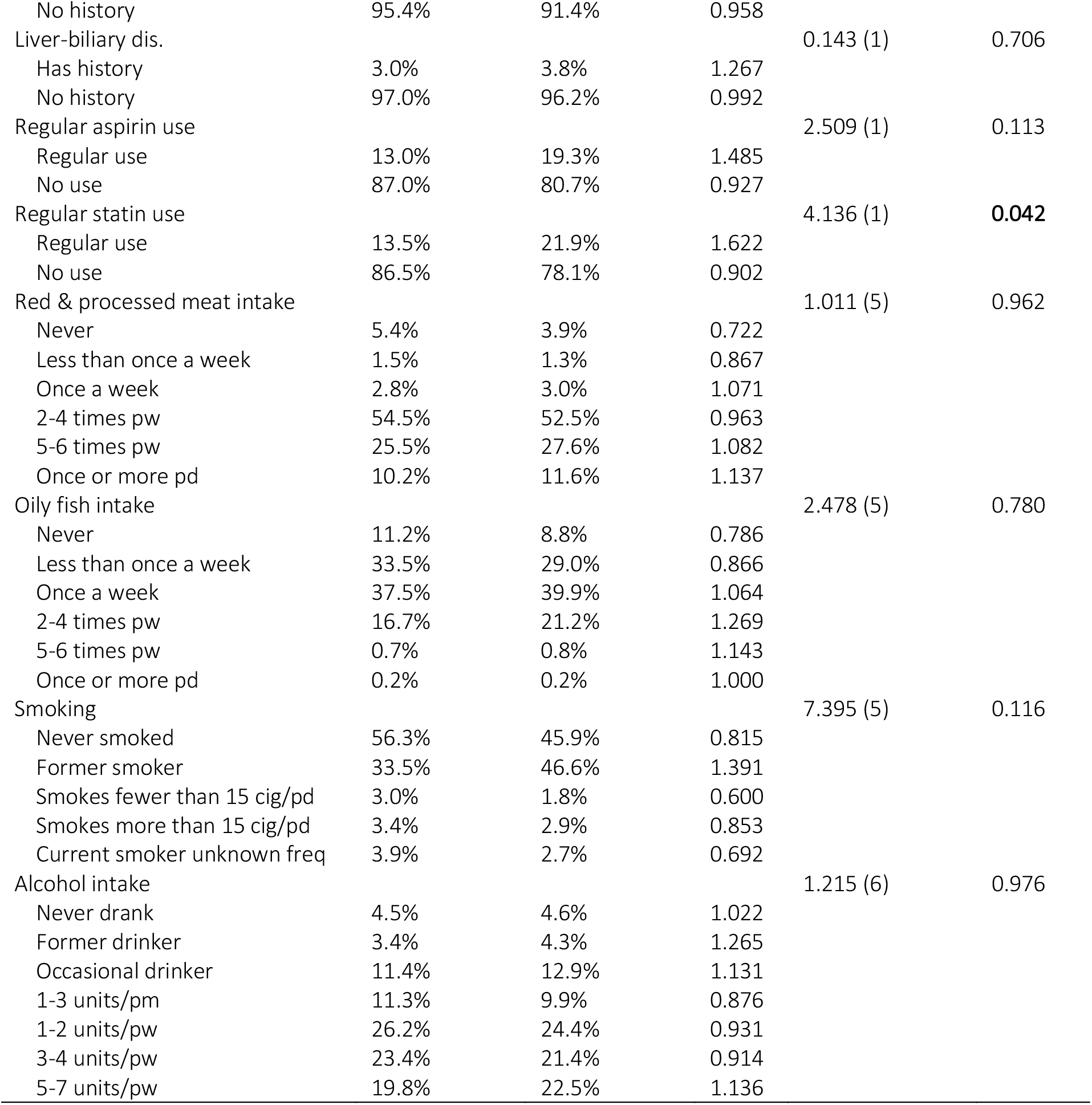
Sample characteristics of the baseline data displaying the group means for continuous measures, distributions, and odds ratios of categories for categorical and ordinal measures. Significant p-values are displayed in bold. Odds ratios above 1 indicate higher incidence in the CRC group compared to the control group. CRC: Colorectal cancer; CSE: Certificate of Secondary Education; CVD: cardiovascular disease; GCSE: General Certificate of Secondary Education; HNC: Higher National Certificate; HND: Higher National Diploma; IBD: inflammatory bowel disease, DI: deprivation index; NVQ: National Vocational Qualification; OR: Odds ratio.

Two groups further differed in their disease history. Compared to controls, the CRC group had a significantly greater odds of having a family history for cancer and CRC and disease history of CVD. This effect was especially high for the family history of CRC, where incidence in CRC group was twice the incidence in controls. The CRC group also had a higher proportion of individuals who have been previously screened for CRC. CRC patients rated their health significantly lower on the self-reported health rating compared to controls. Groups did not show any difference in their diet and lifestyle measurements. There was no significant effect of the study centre (*X*^*2*^(21) = 2.824; *P =* 1.000).

We further explored cancer-related measures in the CRC group. Mean age of diagnosis was 55.9 years (*SD =* 8.04) with a negatively skewed distribution. Throughout the duration of the cancer registry data, 65.9% of CRC group had one, 25.4% had two and 9% had three or more distinct cancer diagnoses. For almost all (99.8%), the tumour was of malignant type in the primary cancer site. Among CRC patients, 4.79% had also developed breast cancer, 2.11% lung cancer, 1.24% non-Hodgkin’s lymphoma, 1.01% prostate cancer and 0.29% liver cancer.

Finally, we compared the physical and biochemical biomarkers across groups. All continuous biomarkers, except for RBC, nucleated RBC, ALT, diastolic BP, IGF-1, grip strength, vitamin D, haematocrit, apolipoprotein A and B, calcium and haemoglobin were significant at the corrected level (*P*_cor_ < 0.05). Detailed results are given in **Supplementary Fig. S1**.

### Cox proportional hazard model

In the initial feature selection step, we removed the following features which were above the *P <* 0.10 threshold: Townsend deprivation score, ethnicity, platelets, apolipoprotein B, low density lipoprotein, total protein, systolic BP, statin use, disease history for CVD, IBD, liver-biliary disease and diabetes, education level, sleep duration, mean corpuscular volume, neutrophils, eosinophils, nucleated RBCs, albumin, ALP, AST, glucose, HbA1C, and phosphate. **Fig 2** shows the hazard ratios and the clustering of the features that remained after the feature selection step. Next, we removed intercorrelated variables that showed a *VIF* > 10. These variables were hematocrit percentage, direct bilirubin, apolipoprotein A, testosterone, impedance, metabolic rate, and height.

**Fig 2.**
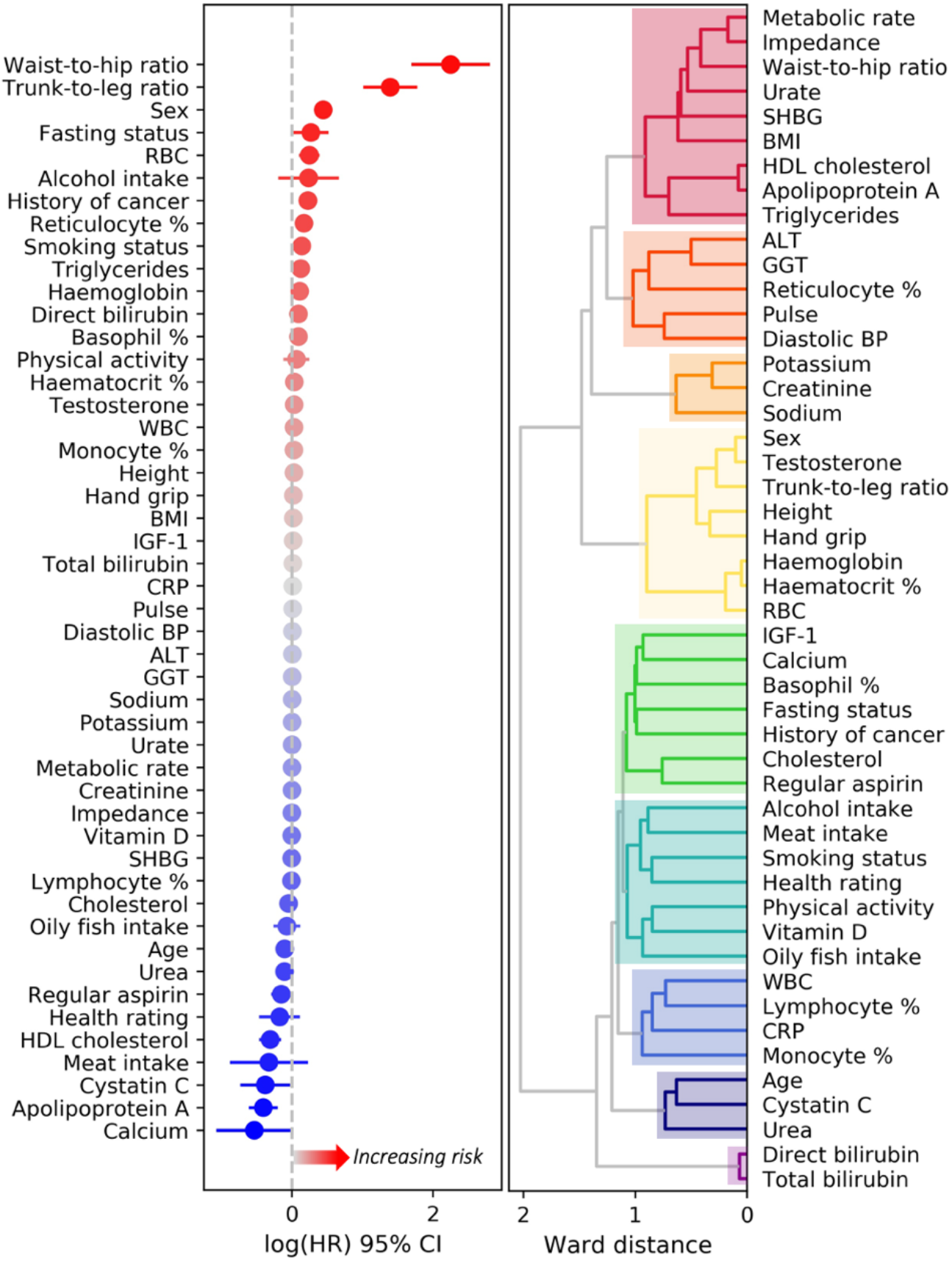
Risk profile and relationships between features that remained in the set after the initial forward selection step. Plot on the left shows univariate log hazard ratios and standard error of each variable. Log hazard ratios above 0 indicate an increased risk for CRC. Whereas values below 0 indicate a decreased risk (i.e., protective effect) for CRC. Dendrogram on the right displays the relationships between variables calculated as the Ward distance of the pairwise intercorrelations. The distance reflects the similarity between variables, where smaller the distance more similar the variables are. For instance, direct bilirubin and total bilirubin are very similar and form their own cluster. From top to bottom biomarkers formed the adiposity & cholesterol (red), hepatic & cardiovascular (dark orange), urine (orange), sex & RBC (yellow), cancer promoting factors & cholesterol (green), lifestyle (teal), immune system (blue), renal function (navy), and liver-biliary cluster (purple).

In the final backward elimination step, we removed variables not significantly contributing to the model: RBC count, grip strength, calcium, trunk-to-leg fat ratio, sodium, creatinine and potassium in urine, lymphocytes, cystatin C, urate, self-reported health rating, high density lipoprotein, oily fish and red meat intake, WBC count, monocytes, hemoglobin, total bilirubin, fasting status, diastolic BP, vitamin D, reticulocytes, CRP, MET minutes, GGT, IGF-1, BMI, regular aspirin use, and smoking. With backward elimination, we reduced the partial Akaike information criterion of the model from 39,044.14 to 39,006.05, whereas the C-index only marginally reduced from 0.696 to 0.690.

The final model (**Fig 3A**) included the following risk factors as strongest predictors for developing CRC: higher waist-to-hip ratio (491% increase), regular alcohol intake (63% increase), being male (32% increase) (**Fig 3B**), and having family history of cancer (29% increase) (**Table 2**). Having higher basophil percentage (10% increase), higher triglycerides (8% increase), higher pulse (1% increase), and higher SHBG (0.3% increase) increased the risk for CRC. Whereas having higher urea (7% decrease), higher total cholesterol (6% decrease), and higher ALT (0.4% decrease) decreased the risk for CRC, suggesting that these might be protective factors against the disease. Younger age at recruitment was associated with a higher risk for CRC, where age at recruitment and at diagnosis showed a high positive correlation (*r* = 0.90). The final model’s C-indices were 0.690 and 0.681 for the training and test sets respectively. As this final model included serum lipids, we ran the model again by including the fasting status. However, the fasting status was not significant and did not affect the results of other predictors. For completeness and comparison in the **Supplementary Table S2** we report the unadjusted hazard ratios when the predictors of the final model were fitted to the data in a univariate fashion.

**Fig. 3.**
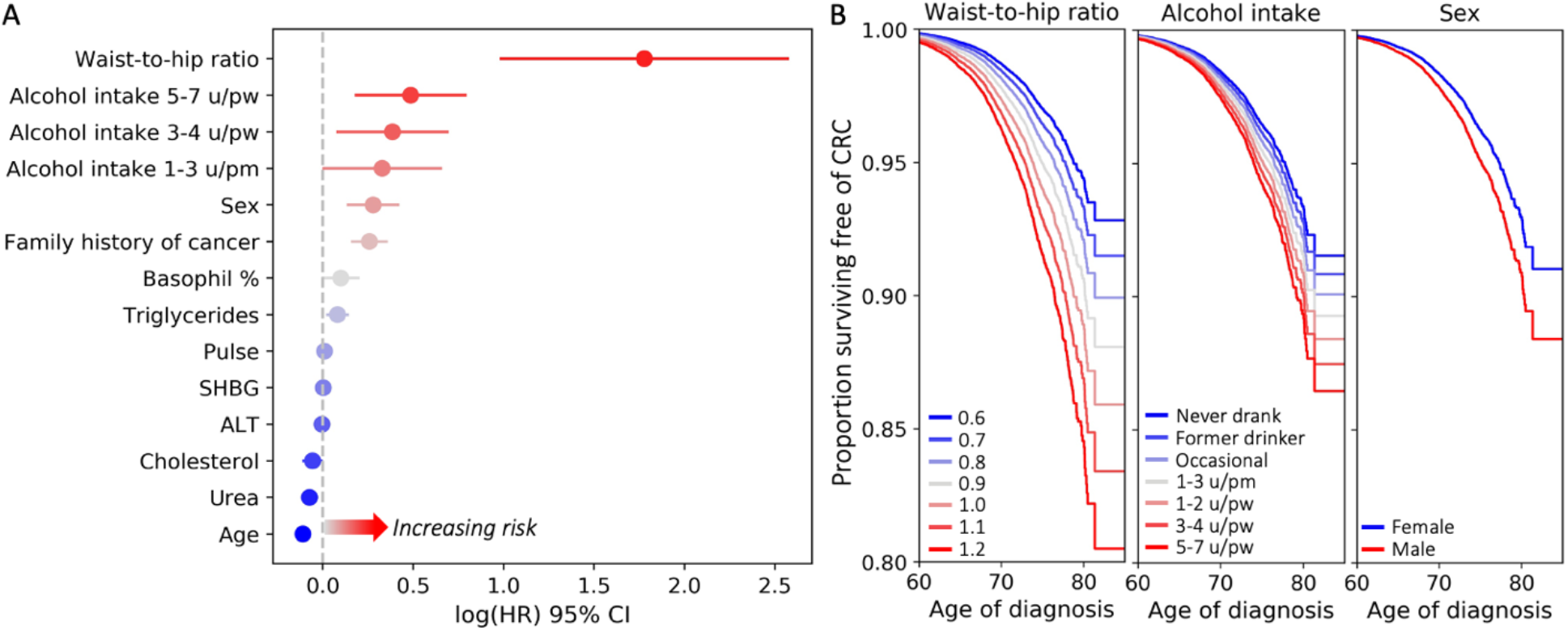
**A**. Results of time-to-event analysis showing log hazard ratios and standard error of each variable included in the final optimal model. Log hazard ratios above 0 indicate an increased risk for CRC. Whereas values below 0 indicate a decreased risk (i.e., protective effect) for CRC. **B**. Partial effects plots for the top 3 risk factors given the model, y axis displaying the proportion of participants who did not develop CRC. Note that the rate of decline in proportion of healthy individuals is faster for participants with waist-to-hip ratio of 1.2, those who drink 5-7 units of alcohol per week and males. CI: confidence interval; HR: hazard ratio; pm: per month; pw: per week; u: units.

**Table 2.** Results of the final Cox PH model in the descending order of the HR magnitude. Columns display the predictor name, the mean or percentage of the predictor for each group, hazard ratio, confidence interval and p value respectively. HRs higher than 1 indicate increase in risk for CRC, whereas HRs lower than 1 indicate a decrease in risk. CI: confidence interval; HR: hazard ratio; ref: reference category; SHBG: sex hormone binding globulin; u: units.

| Predictor | Controls | CRC | HR | 95% CI | P |
| --- | --- | --- | --- | --- | --- |
| Waist-to-hip ratio | 0.88 ± 0.09 | 0.91 ± 0.09 | 5.917 | 2.681 – 13.060 | **< 0.00005** |
| Alcohol intake (ref: Never drank): |  |  |  |  |  |
| Former drinker | 2.97% | 2.54% | 1.082 | 0.720 – 1.625 | 0.705 |
| Occasional | 9.92% | 8.49% | 1.148 | 0.826 – 1.595 | 0.412 |
| 1-3 u/pm | 10.66% | 9.61% | 1.388 | 1.005 – 1.919 | **0.047** |
| 1-2 u/pw | 26.55% | 23.29% | 1.254 | 0.926 – 1.699 | 0.143 |
| 3-4 u/pw | 24.85% | 24.98% | 1.470 | 1.087 – 1.989 | **0.012** |
| 5-7 u/pw | 21.22% | 27.89% | 1.626 | 1.203 – 2.198 | **0.002** |
| Unknown | 0.09% | 0.13% | 2.410 | 0.749 – 7.749 | 0.140 |
| Male sex (ref: Female) | 55.21% | 66.83% | 1.321 | 1.151 – 1.515 | **0.0001** |
| Family history of cancer (ref: No history): |  |  |  |  |  |
| True | 33.63% | 40.94% | 1.294 | 1.177 – 1.422 | **< 0.00005** |
| Unknown | 1.87% | 1.74% | 0.893 | 0.614 – 1.302 | 0.840 |
| Age | 55.35 ± 8.18 | 60.18 ± 6.82 | 0.896 | 0.885 – 0.906 | **< 0.00005** |
| Basophil % | 0.55 ± 0.45 | 0.56 ± 0.50 | 1.105 | 1.003 – 1.217 | **0.043** |
| Urea | 5.35 ± 1.25 | 5.47 ± 1.31 | 0.929 | 0.894 – 0.966 | **0.0002** |
| Triglycerides | 1.65 ± 0.91 | 1.79 ± 1.00 | 1.085 | 1.027 – 1.146 | **0.004** |
| Total cholesterol | 5.54 ± 0.98 | 5.48 ± 1.03 | 0.945 | 0.900 – 0.991 | **0.021** |
| Pulse | 68.30 ± 10.88 | 69.57 ± 11.28 | 1.010 | 1.005 – 1.014 | **< 0.00005** |
| ALT | 23.82 ± 12.72 | 23.96 ± 12.18 | 0.996 | 0.991 – 1.000 | **0.041** |
| SHBG | 49.16 ± 25.10 | 47.91 ± 23.32 | 1.003 | 1.000 – 1.005 | **0.030** |

### Classification analysis

In 5-fold CV experiments with RFE, N_features_ of 5 achieved the highest mean AUC score of 0.744. The mean AUC decreased as the number of selected features decreased (0.742 for N_features_=4, 0.734 for N_features_=3, 0.717 for N_features_=2, and 0.690 for N_features_=1) **(Fig. 4B)**. Feature ranking by gain-based importance for the top 20 features is shown in **Fig. 4A** for the 5-fold CV experiments. On average, age had the highest importance score (*M* = 0.354; *SD* = 0.014), followed by self-reported overall health rating (*M* = 0.094; *SD* = 0.010), ALP (*M =* 0.043; *SD =* 0.007), WBC count (*M* = 0.041; *SD* = 0.012), lymphocyte percentage (*M* = 0.040; *SD* = 0.017), family history of cancer (*M =* 0.028; *SD* = 0.011), urine creatinine (*M* = 0.023; *SD* = 0.014), trunk-to-leg ratio (*M =* 0.022; *SD =* 0.007), urine potassium (*M =* 0.018; *SD* = 0.005), and platelet count (*M* = 0.016; *SD* = 0.012). Hence, age, self-reported health rating, ALP, WBC count, and lymphocyte percentage were selected as the final feature set.

**Fig. 4.**
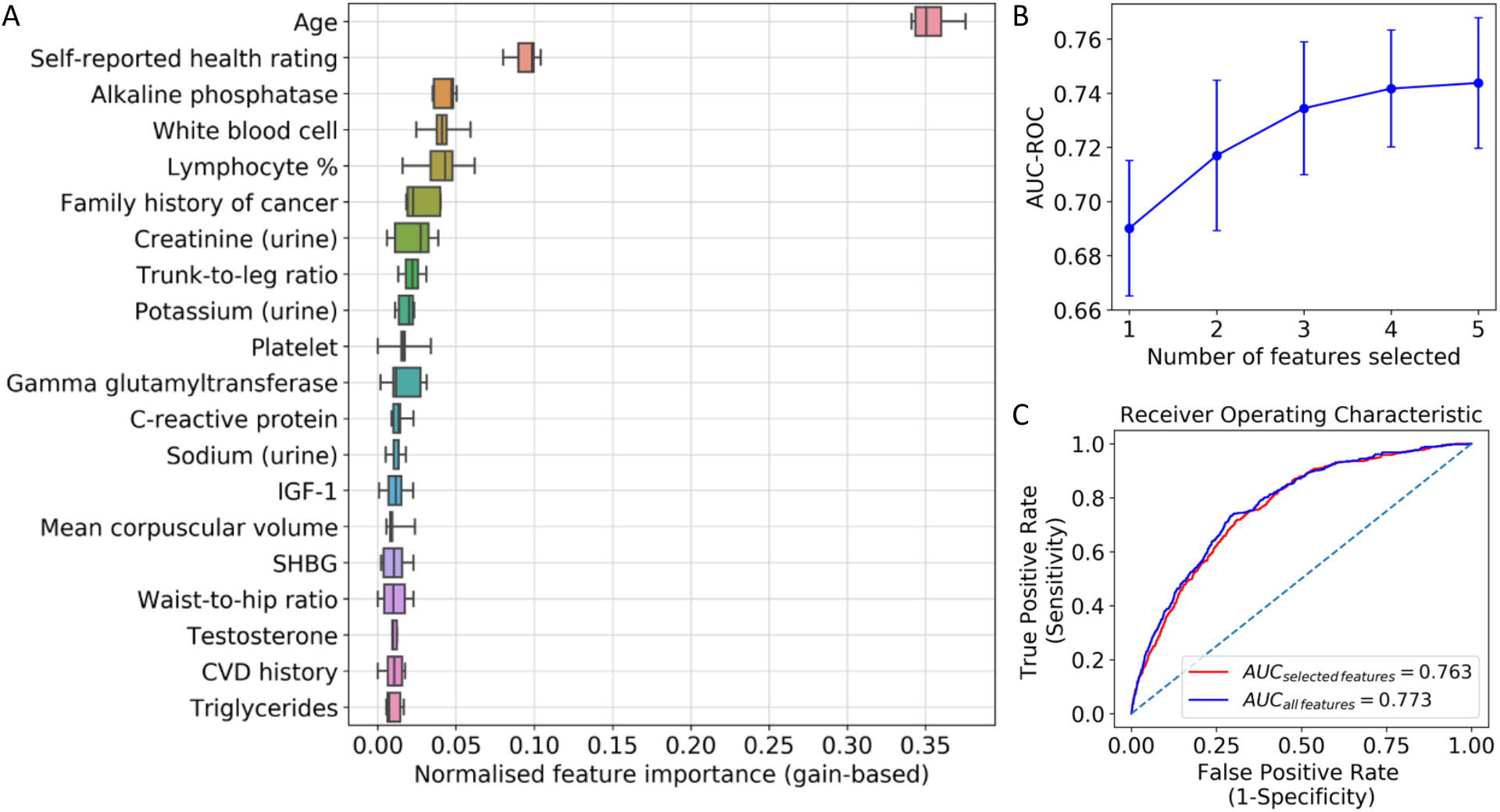
Classification results. **A**. Feature ranking by gain-based importance obtained from tree-boosting random effects model, showing top 20 features. Feature importance values were calculated for each split of the 5-fold cross- validation. For each feature, a boxplot is provided showing distribution of normalised importance across runs. **B**. 5- fold cross-validation results in AUC-ROC for tree-boosting model with number of selected features from 1 to 5 using Recursive Feature Elimination (RFE). **C**. ROC curve for tree-boosting random effects model on the test set with AUC score.

The ROC curve is provided in **Fig. 4C** for the final tree-boosting with random effects model refitted on the whole training set, using only the selected combination of features as well as all available features. The model achieved an AUC score of 0.763 and 0.773 respectively on the test set. At the threshold maximizing sensitivity and specificity equally, the model with the selected combination of features achieved a sensitivity of 0.718 and a specificity of 0.690. PDPs showing marginal contribution of each feature on CRC probability are provided in **Supplementary Fig. S2**.

## Discussion

In this study, we opted to take a two-pronged, data-driven approach to identify pre-diagnostic and diagnostic biomarkers for detecting CRC. The principal result of our study is that in addition to modifiable lifestyle factors of obesity, alcohol consumption and cardiovascular health, blood-based biomarkers that capture WBC activity, lipid profile, liver and kidney function are informative of CRC, at both pre-clinical and clinical stages.

Among blood-based biomarkers we found those measuring lipid metabolism, liver function and immune response to be also sensitive to CRC. We showed that a lipid profile of high triglycerides and low cholesterol to be indicative of diagnostic CRC. Elevated triglycerides have been previously linked to CRC [25], whereas the exact role of cholesterol in CRC is contested. Some studies report a positive association between serum cholesterol and CRC risk [26] and beneficial effect of long-term statin use [27]. Supporting current findings, others reported that low cholesterol increases the risk [17], almost by 2-folds [28]. This inverse relationship can be due to the high energy demanded by cancer cells for proliferation, which is satisfied by increased uptake of the exogenous cholesterol [29], or alternatively by deregulations of its metabolic pathway due to genetic alterations [30].

Imbalances in ALT and ALP, markers of liver dysfunction, are often linked to non-cancer factors like liver injury due to statin use, obesity, diet [31], high alcohol intake [32], and smoking [33]. However, they also relate to the cancers of digestive organs including CRC [33] and liver metastases [15,16]. Our analysis showed an association between elevated levels of ALP and CRC, particularly above 100 U/L. Although this might imply liver metastasis, only a subset of our CRC cases had liver cancer, suggesting that liver markers may have a diagnostic value in the early stages of CRC. Further, the survival analysis highlighted a reduction in ALT in CRC. Lower ALT is linked to oxidative stress [34], having invasive cancer cells [35], poorer prognosis and mortality [36]. Therefore, ALP/ALT ratio can be an informative liver function marker in CRC.

Among the immunity markers, we observed strong associations for basophil and lymphocyte percentage and WBC count. Basophils showed a modest, positive link to CRC risk, which might suggest a diagnostic inflammatory response to cancer [37]. Lymphocytes play an important role in body’s immune response against cancerous cells and lower levels were found to be associated with greater likelihood of CRC [38]. However, the negative association we observed for WBC differed from previous findings [13,14]. This could be explained by chemotherapy-related WBC reduction, previously reported for multiple cancers including CRC [39].

Waist-to-hip ratio and alcohol intake were the strongest modifiable lifestyle risk factors, with one unit increase in waist-to-hip ratio increasing the CRC risk by nearly 500%. The association is supported by previous studies [40], and retained even when the model was adjusted for BMI, confirming that the association is due to visceral fat but not obesity [41]. High alcohol intake leads both to higher visceral fat and CRC [42]. We found that compared to non-drinkers, moderate-to-heavy drinking increased the hazard by 62% in a dose-dependent fashion, such that highest risk was observed for highest consumers. Although not significant, there were a higher percentage of former and moderate-to-heavy drinkers in the CRC group. Concordantly, previous work suggested that while moderate and heavy drinking was associated with increased risk, light consumption was not [43]. Visceral adiposity and high alcohol intake may promote carcinogenesis via hyperinsulinemia, elevated oxidative stress and inflammation pathways [44].

Sex is a prominent determinant in CRC development. Here we report over 30% increased risk in men. There are more men who develop CRC than women, 84.5 versus 56.5 per 100,000, and higher mortality in men in the UK [3]. This susceptibility may be due to behavioural and biological factors [45]. Men have a higher red meat intake [46], alcohol consumption [47], and tendency to deposit visceral fat [48], all being significant risk factors. Sex hormones might also contribute to the sex-related differences in risk. We found a modest positive association between CRC risk and SHBG, a glycoprotein responsible for transporting sex hormones estrogen and testosterone. Higher levels of SHBG would lead to lower levels of circulating free testosterone and estrogen which are protective against CRC [49].

Finally, we report family history of cancer, higher pulse, lower urea and poor health rating as other significant contributors to CRC risk. History of cancer amplified CRC risk by 29%. CRC group had a higher incidence of family history with CRC (OR = 2.03) and cancer (OR = 1.34). First degree relatives of CRC patients have a higher risk for overall cancer, mainly CRC, thyroid, and stomach [50]. The risk of getting colon cancer in a first-degree relative was 2.2, which drops to 1.3 and 1.2 in second and third-degree relatives [51], suggesting a strong familial genetic component. Whereas pulse and urea had minor contributions to the risk, emphasizing the modest but key role of cardiovascular [52] and renal health [53] in CRC development [53]. Finally, self-reported poor health was a strong classifying feature, previously linked to overall cancer risk and all-cause mortality [54,55]. While these biomarkers may not be necessarily specific to CRC on their own, when combined they can yield a higher specificity.

ML models can detect subtle changes and non-linear relationships in biomarkers, which may not be directly evident to clinicians. However, only a limited number of ML studies have investigated the use of routinely collected, longitudinal clinical data in detecting CRC. There is notable work that extracted blood test results from electronic healthcare records and used ML methods to identify patients at risk of CRC or with CRC [56,57]. The former study used a predictive algorithm involving an ensemble of random forest and gradient boosting to estimate the risk of developing CRC (AUC=0.82). The latter evaluated five ML models, including logistic regression, naïve Bayes and support vector machines, alongside feature selection to identify the best model and features for distinguishing CRC from healthy (AUC=0.85).

Similarly, in the current study, we show the utility of ML applications in healthcare, and their potential to improve diagnostics when combined with other powerful statistical methods.

CRC has an immense economic burden on the healthcare system. Direct and indirect costs in the UK constitute £314m and £1.4bn respectively [58]. These costs can be drastically reduced with the help of improved diagnostic tools, and increased uptake of screening programmes. Current screening in the UK involves FIT (£4 per kit) [59], and a follow-up colonoscopy (£465 pp) [60]. Reducing the false negative rate of FIT by combining it with blood-based biomarkers sensitive to CRC, may increase its diagnostic accuracy and help reduce the long-term economic burden. Blood-based biomarkers offer a key advantage: they are routinely collected laboratory tests, and their results are available at low to no additional cost. This allows clinicians to interpret the FIT with readily available blood test results, and subsequently stratify and triage patients.

Our study had several strengths and limitations. We have opted to take a two-pronged data-driven approach that used survival analysis to determine the pre-diagnostic variables associated with developing CRC throughout the course of the study, and tree-boosting model to determine the clinical features that can classify CRC. We opted for methods that maximally utilize the available data and tested all available variables in our dataset. In order to find the optimal combination of biomarkers both in the survival analysis and in the GPBoost model, we adopted feature selection methods widely used in ML. The study and analyses were planned retrospectively based on the data and participants available in the UK Biobank study. Due to the large sample size of the study, CRC group was well-represented. However, our analyses were limited by the measurements collected in the study which were not specific to cancer. Biochemistry measures did not include other widely used diagnostic measures of cancer such as CEA and FIT. Therefore, we are not able to quantify the additive value of the blood-based biomarkers we report in this study to other CRC screening tools.

In conclusion, in the current study, we used Cox regression to identify key biomarkers that can predict CRC risk from baseline measurements and thus can be used for early detection of disease, and a tree-boosting classification model to identify biomarkers that can distinguish between healthy and CRC states. We provide novel evidence that blood-based biomarkers collected in routine blood examinations, capturing immune response, lipid profile, liver and kidney function are informative of preclinical and clinical CRC. These blood-based biomarkers can provide an additive value to the current, widely used CRC diagnostic tools, to help improve their diagnostic accuracy, increase uptake, and allow earlier disease detection.

## Data Availability

Approval for the study and permission to access the data was granted by the UK Biobank Resource. UK Biobank is an open access resource and bona fide researchers can access the UK Biobank dataset by registering and applying at https://www.ukbiobank.ac.uk/enable-your-research/apply-for-access. Code for the analysis will be made available on GitHub at https://github.com/sanometech/ukbiobank-crc/.

## Abbreviations

ALP: alkaline phosphatase
ALT: alanine aminotransferase
AST: aspartate aminotransferase
AUC: area under curve
BMI: body mass index
BP: blood pressure
C-index: concordance index
CEA: carcinoembryonic antigen
CRC: colorectal cancer
CV: cross-validation
CVD: cardiovascular disease
FBC: full blood count
FIT: faecal immunochemical test
HbA1C: glycated haemoglobin
HC: healthy controls
HR: hazard ratio
IBD: inflammatory bowel disease
ICD-10: International Classification of Diseases 10th revision
GGT: gamma glutamyltransferase
IGF-1: insulin-like growth factor 1
MET: metabolic equivalent task
ML: machine learning
OR: odds ratio
PDP: partial dependence plots
pd: per day
pm: per month
pw: per week
RFE: recursive feature elimination
ROC: receiver operating characteristic curve
SHBG: sex hormone binding globulin
u: unit
VIF: variance inflation factor
WBC: white blood cells.

## Acknowledgements

This research has been conducted using the UK Biobank Resource under application number 87991 for the project titled ‘Validation of an AI-powered online search strategy for finding optimal biomarker combinations’. We thank the UK Biobank participants and researchers who built the UK Biobank Resource.

## Authors’ contributions

G.T. and E.K. had full access to the dataset as the primary investigator and collaborator respectively. Study conception and design: G.T. Statistical analysis: G.T. and E.K. Interpretation of results: G.T. and E.K. Writing manuscript: G.T. and E.K.

## Competing interest

The authors declare no competing interests.

## Funding information

UK Biobank study was primarily funded by the Wellcome Trust and the Medical Research Council. The present work received no external funding.

## Notes

### Competing Interest Statement

The authors have declared no competing interest.

### Author Declarations

Approval for the study and permission to access the data was granted by the UK Biobank Resource, under application number 87991 for the project titled Validation of an AI-powered online search strategy for finding optimal biomarker combinations.

