## Supplementary information for "Additive pre-diagnostic and diagnostic value of routine blood-based biomarkers in the detection of colorectal cancer in the UK Biobank cohort"

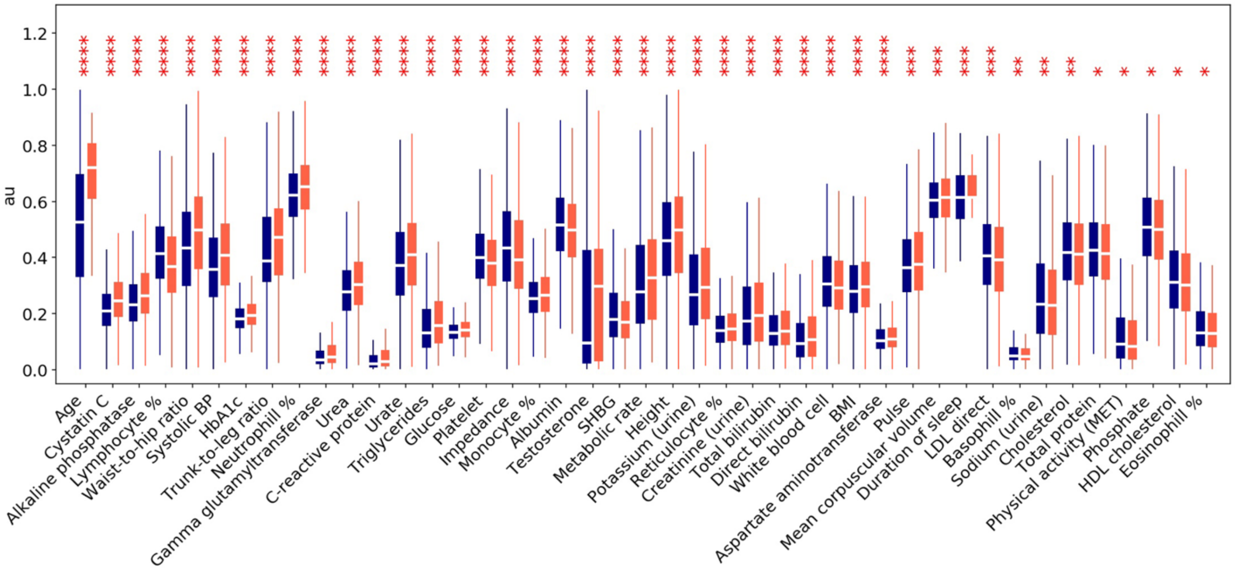


**Fig S1:** The distribution of continuous biomarkers that showed significant group differences at the corrected level. Blue and red boxes represent HC and CRC group respectively. Boxplots are displayed in descending order of effects from left to right with significance. The data are rescaled between 0-1 within each variable, to allow comparison across measures. Au: arbitrary units. **** $P < 0.0001$, *** $P < 0.001$, ** $P < 0.01$, * $P < 0.05$.

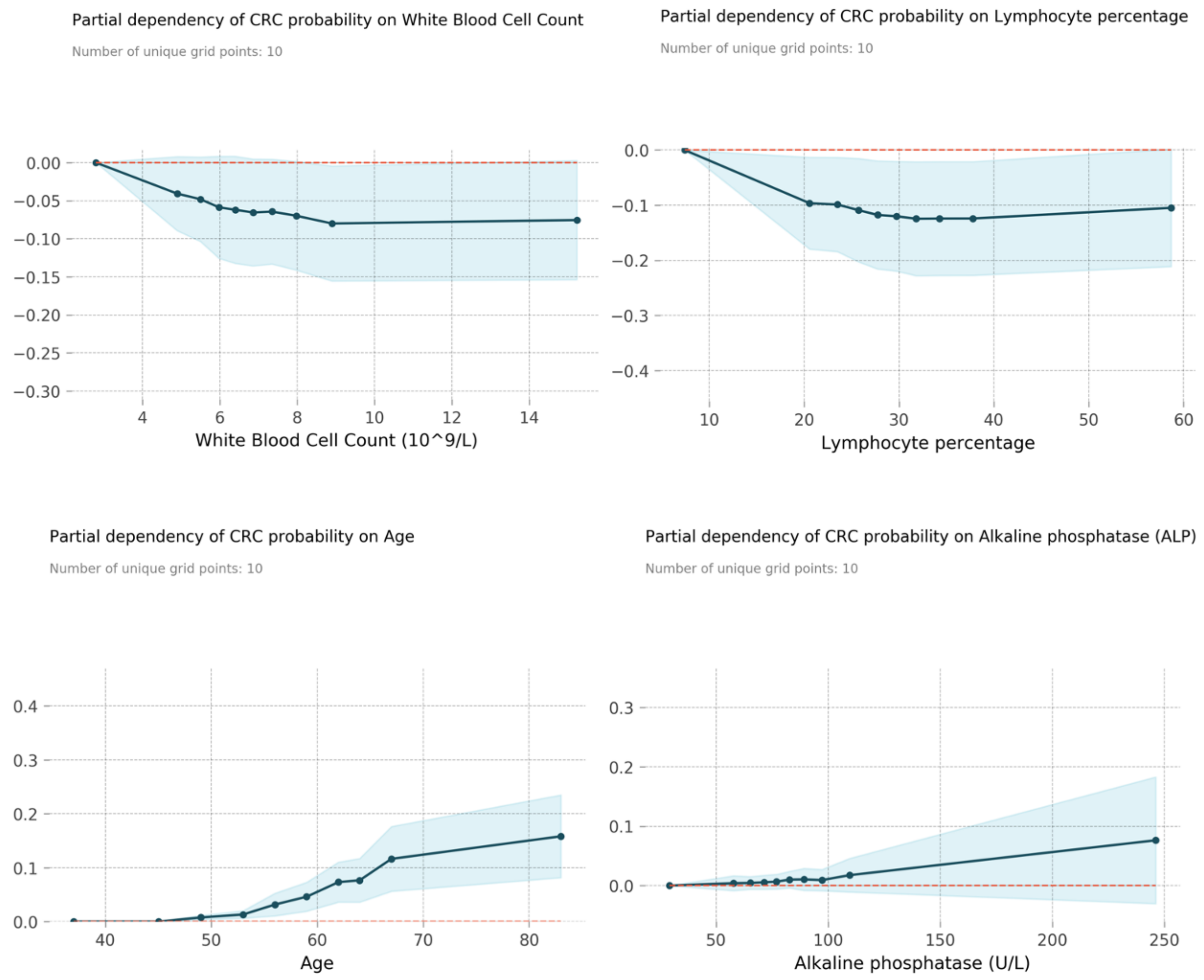


**Fig. S2:** Partial dependency plots showing marginal contribution of each feature included in the final classification model. Blue lines show the average contribution of feature on the CRC probability and shaded areas represent level of confidence.

**Table. S1:** Univariate (unadjusted) results of the predictors in the final Cox PH model. Columns display the predictor name, hazard ratio, confidence interval, p value and the C-index respectively. HRs higher than 1 indicate increase in risk for CRC, whereas HRs lower than 1 indicate a decrease in risk. Higher C-indices indicate better model fit. CI: confidence interval; C-index: concordance index; HR: hazard ratio; ref: reference category; SHBG: sex hormone binding globulin; u: units.

| Predictor | *HR* | 95% *CI* | *P* | C-index |
|---|---|---|---|---|
| Waist-to-hip ratio | 9.485 | 5.555 – 16.195 | **<0.0005** | 0.545 |
| Alcohol intake (ref: Never drank): | | | | 0.540 |
| Former drinker | 1.265 | 0.842 – 1.899 | 0.257 | |
| Occasional | 1.175 | 0.846 – 1.633 | 0.337 | |
| 1-3 u/pm | 1.545 | 1.119 – 2.134 | **0.008** | |
| 1-2 u/pw | 1.420 | 1.050 – 1.921 | **0.023** | |
| 3-4 u/pw | 1.692 | 1.253 – 2.284 | **0.001** | |
| 5-7 u/pw | 1.820 | 1.350 – 2.454 | **<0.0005** | |
| Unknown | 3.009 | 0.936 – 9.666 | 0.064 | |
| Male sex (ref: Female) | 1.557 | 1.411 – 1.718 | **<0.0005** | 0.544 |
| Family history of cancer (ref: No history): | | | | 0.524 |
| True | 1.253 | 1.140 – 1.377 | **<0.0005** | |
| Unknown | 0.900 | 0.618 – 1.311 | 0.583 | |
| Age | 0.900 | 0.890 – 0.910 | **<0.0005** | 0.644 |
| Basophil % | 1.096 | 0.995 – 1.208 | 0.063 | 0.498 |
| Urea | 0.898 | 0.865 – 0.933 | **<0.0005** | 0.552 |
| Triglycerides | 1.132 | 1.079 – 1.188 | **<0.0005** | 0.511 |
| Total cholesterol | 0.952 | 0.911 – 0.996 | **0.032** | 0.512 |
| Pulse | 1.010 | 1.006 – 1.014 | **<0.0005** | 0.540 |
| ALT | 1.006 | 1.002 – 1.009 | **0.003** | 0.512 |
| SHBG | 0.995 | 0.993 – 0.997 | **<0.0005** | 0.537 |
